# Early detection of physiological deterioration in post-surgical patients using wearable technology combined with an integrated monitoring system: a pre- and post-interventional study

**DOI:** 10.1101/2020.12.01.20240770

**Authors:** Peter J Watkinson, Marco AF Pimentel, Lei Clifton, David A Clifton, Sarah Vollam, Duncan Young, Lionel Tarassenko

## Abstract

**Objectives:** Late recognition of physiological deterioration is a frequent problem in hospital wards. We assessed whether ambulatory (wearable) physiological monitoring combined with a system that continuously merges physiological variables into a single “risk” score (VSI), changed care and outcome in patients after major surgery.

**Design:** Pre- and post-interventional study.

**Setting:** A single centre tertiary referral university hospital upper-gastrointestinal service.

**Participants:** Patients who underwent major upper-gastrointestinal surgery.

**Interventions:** Phase-I (pre-intervention phase): Patients received continuous wearable monitoring and standard care, but the VSI score was not available for clinical use. Phase-II (post-intervention phase): Patients received continuous wearable monitoring. In addition to standard care the VSI score was displayed for use in clinical practice.

**Measurements and Main Results:** 200 participants were monitored in phase-I. 207 participants were monitored in phase-II. Participants were monitored (median, interquartile range, IQR) for 30.2% (13.8-49.2) of available time in phase-I and 58.2% (33.1-75.2) of available time in phase-II.

Clinical staff recorded observations more frequently in the 36 hours prior to a major adverse event (death, cardiac arrest or unplanned admission to intensive care) for phase-II participants (median, IQR, time between observations of 1.00, 0.50-2.08 hours) than phase-I participants (1.50, 0.75-2.50 hours, *p*<0.001). There was no difference in observation frequency between the two phases for participants who did not undergo an adverse event (*p*=0.129). 6/200 participants died before hospital discharge in phase-I, 1/207 participants died in hospital in phase-II. 20 (10.0%) patients in phase-I and 26 (12.6%) patients in phase-II had an unplanned admission to intensive care. Ward length-of-stay was unaltered (8.91, 6.71-14.02 days in phase-I, vs. 8.97, 5.99-13.85 days in phase-II, *p*=0.327).

**Conclusion:** The combination of the integrated monitoring system with ambulatory monitoring in high-risk post-surgical patients improved recognition and management of deteriorating patients without increasing the observation rate in those patients who did not deteriorate.

## INTRODUCTION

Deterioration in hospital after major surgery has long-term outcome effects even if the participant survives the acute episode (1). Delayed recognition of deterioration has been associated with substantially worse outcomes (2,3,4). The UK National Health Service (NHS) database of patient safety incidents recorded over 2000 preventable deaths in adult NHS patients in England between 2010 and 2012 (5). Analysis of these incidents involving inpatient mortality revealed that the most common factor was mismanagement of patient deterioration, which accounted for 35% of these deaths. This included failure to act on or recognise deterioration, failure to give ordered treatment or support in a timely manner, and failure to observe patients’ vital signs.

Changes in vital signs (blood pressure, heart rate, respiratory rate, temperature, and oxygen saturation) aid recognition of deterioration (6,7). Current early-warning scoring (EWS) systems have poor positive predictive values for events in patients after surgery (8,9). They are limited by the intermittent nature of the observations recorded (10). When a patient deteriorates, it is likely that their vital signs will change between “routine” assessments. Earlier detection may therefore be assisted by continuous monitoring systems. However, patients do not normally have their vital signs continuously monitored unless they are either at high risk, or have been recognised as having deteriorated. In ward environments, mobility is encouraged as part of recovery. Wearable monitors are therefore required. Moreover, current EWS systems do not take account of the relationships between changes in individual vital signs. In contrast, the integrated monitoring system (IMS) Visensia (formerly BioSign) takes account of these by modelling their relationship in a 5-dimensional space (11). The performance of the system connected to static monitors was retrospectively assessed in a randomised controlled trial study in high-risk patients outside of critical areas compared with standard ward care (12,13). However, the system has not yet been tested in an ambulatory hospital setting.

We conducted a prospective, before-and-after study in which we assessed the effects of the addition of wearable monitoring combined with the integrated monitoring system in participants following major gastrointestinal surgery.

## MATERIALS AND METHODS

### Study design

We conducted a prospective, before-and-after interventional study to assess the efficacy of wearable monitoring combined with an integrated monitoring system in participants following major surgery (EudraCT No: 2011-000928-15). The study was approved by the local ethics committee (Oxford Research Ethics Committee OxREC No: 08/H0607/79).

### Setting

We did the study on the upper-gastrointestinal surgical ward in Oxford University Hospitals NHS Trust.

### Participants

We identified potential participants by screening pre-operative assessment bookings and operation lists. Participants were eligible to take part in the study if they were planned to undergo major upper-gastrointestinal surgery. We defined major upper-gastrointestinal surgery as oesophagectomy, oesophagogastrectomy, gastrectomy, Whipples operation, liver resection, pancreatectomy, gastric bypass, biliary reconstruction and splenectomy. Exclusion criteria for the study are detailed in Table-A1 (Supplemental Digital Content). We approached participants either in the pre-operative assessment clinic or on admission.

### Equipment and procedures

Clinical staff received training by research nurses in the use of study equipment before the start of each phase. Participants were followed up daily by research staff during the working week and once over the weekend. Ward staff connected participants to a standard ward bedside monitor (Phillips M3046A/Intellivue MP50, www.healthcare.philips.co.uk) when they arrived on the ward after surgery. The bedside monitors were connected to an external tablet device (Toughbook Getac CA27, uk.getac.com). The tablet allowed the bedside vital-sign observations to be transmitted via the hospital wireless network to a secure server. Clinical staff transferred participants to ambulatory monitoring when they considered a participant safe to be disconnected from “static” bedside monitoring. Participants wore ambulatory monitoring until they were deemed fit to discharge by the clinical team, or the participant requested to stop wearing the kit. Ambulatory monitors consisted of wearable pulse oximeters (Avant-4100 Wrist-Worn, Nonin, www.nonin.com) and electrocardiograph monitors (Corscience-BT3/6 wireless PC electrocardiogram, www.corscience.de/en) connected via Bluetooth (v.2) to enabled PDAs (HTC Touch 3G devices – no SIM, www.htc.com/uk). We stopped using the Corscience electrocardiogram devices after 25 patients as they were very poorly tolerated. When a participant wore ambulatory monitoring, we requested ward nurses to input respiratory rate, temperature and blood pressure manually into the enabled PDAs when they did their routine observations. We transferred pulse oximetry and vital-sign observations entered by clinical staff from the PDA via the hospital wi-fi to a central server. We installed the integrated monitoring system on this server. The Visensia IMS is a U.S. Food and Drug Administration-approved patient monitoring system. It uses a probabilistic model derived from the five vital signs (heart rate, respiratory rate, blood pressure, temperature, oxygen saturation) collected from a representative group of at-risk participants to produce a single numerical score, the VSI (11). The VSI is displayed as a number from 0.0 (no physiological derangement) to 5.0 (severe physiological derangement). An alert is generated only when VSI exceeds a threshold of 3.0 for more than four of the preceding five minutes (11). VSI is updated each time new data are received. If a vital sign is not available for over an hour the mean value from the model for that vital sign is used to calculate VSI.

### Study Phases

The study was conducted in two phases. In phase-I, the IMS was active but not displayed to attending staff; we recorded the continuously-monitored vital signs along with the (non-visible) VSI. Patients received standard ward care, including established use of a conventional paper-based EWS system (Supplemental Digital Content – Table-A2). Completeness of both bedside and ambulatory monitoring data was reviewed every two weeks throughout phase-I, which allowed the development of standard operating procedures for phase-II in order to maximize the amount of time for which patients were monitored.

Prior to commencing phase-II, phase-I data were reviewed to assess the performance of the VSI with ambulatory monitoring in the upper-gastrointestinal surgical population. To minimise time without monitoring, we instituted twice-daily visits to the ward by research staff; research staff could also remotely check that monitoring was taking place, and contact the ward to request re-connection.

In phase-II, the VSI was displayed on the tablet by the participant’s bedside, or on the PDA if they had been transferred to ambulatory monitoring (Figure 1). The index was also displayed, together with the participants’ details, on large touch screens opposite the nurse workstations. Actions following a VSI alert were at the discretion of the ward staff. Intensive care staff from the adjacent Intensive Care Unit (ICU) were available to review patients as requested by ward staff, after their direct evaluation of the participant. As in phase-I, the standard EWS system remained in use, in accordance with normal ward care. Participants were followed to hospital discharge or 30 days after admission to the ward on both phases of the trial.

**Figure 1.**
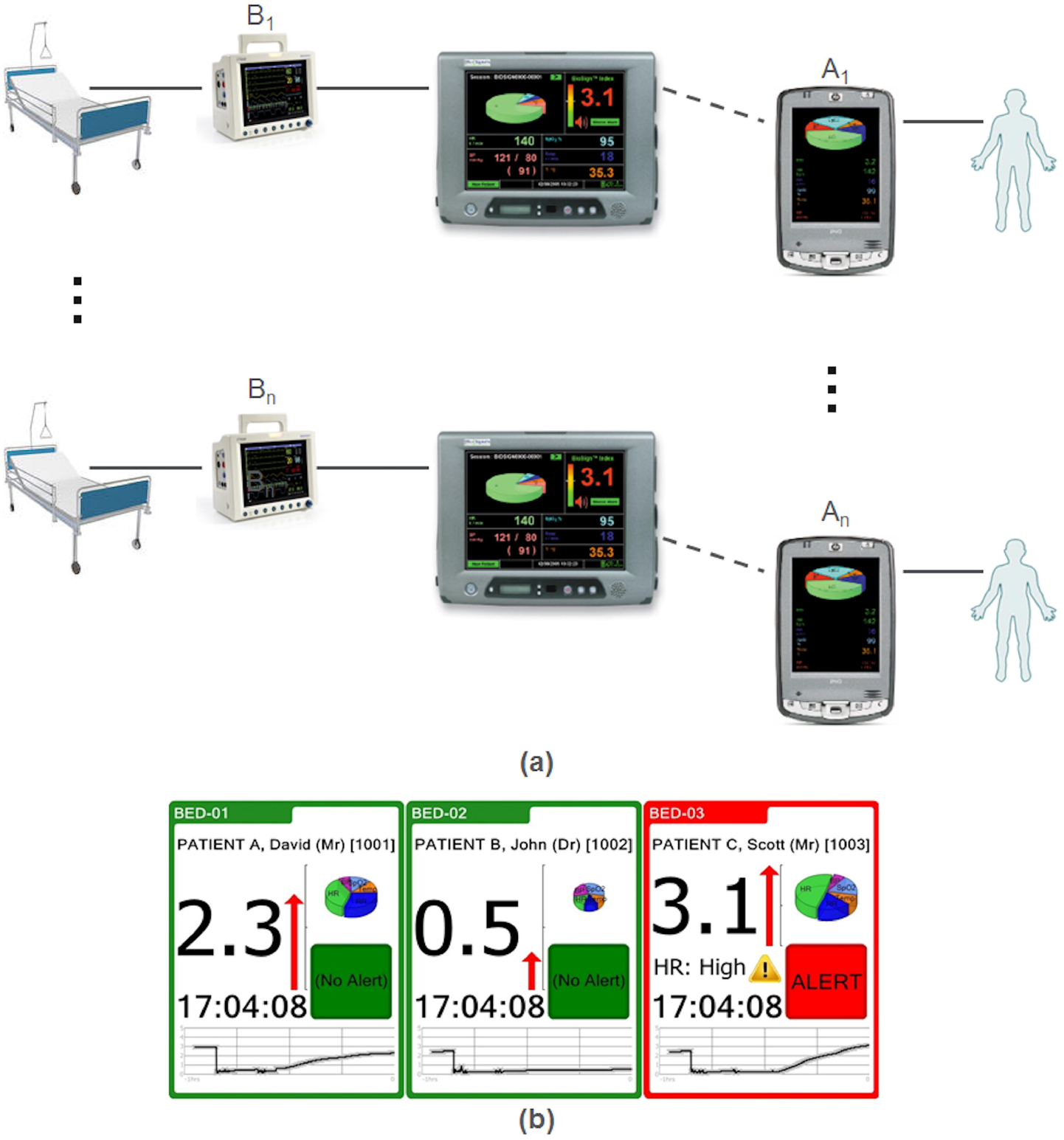
**(a)** Schematic representation of the infrastructure for the study conducted. Patients confined to beds were connected to conventional bedside vital-sign monitors (denoted *B*_*n*_), which in turn are wired to a patient monitor on which the Visensia Integrated Monitoring System (IMS) is implemented. When patients are ambulatory, they have their vital signs measured using mobile sensors or entered manually in the PDA (denoted *A*_*n*_), which are then connected wirelessly (using the hospital network) to the patient monitors. Solid lines denote wired connections and dashed lines denote wireless connections. **(b)** Visensia IMS display for alerting and non-alerting patients. The icons for participants scoring <3.0 were highlighted in green. The icon for participants scoring ≥3.0 flashed red until clinical staff acknowledged the alert on the touch screen (after which it remained red until the score decreased). If no vital signs were available in the previous hour the last calculated index was displayed in grey. Each icon contained a pie-chart showing the degree to which each vital sign was contributing to the score.

### Study size and outcome measures

We hypothesised that if the IMS has a clinically significant effect by appropriate early warning avoiding or decreasing the severity of clinical deteriorations, it should cause a clinically significant reduction in the ward length-of-stay. A sample size of 200 patients per arm (i.e., phase) was required to achieve 80% power to detect a 2-day reduction in the mean hospital length-of-stay (the primary endpoint) at a two-sided significance level of 0.05. Secondary outcome measures included the number of major adverse events (defined as a cardiac arrest, death or unplanned admission to ICU), unplanned returns to the operating theatre, time to ICU admission from the ward, cardiac arrest calls, in-hospital mortality, 30-day mortality post-ward admission, and assessment of the performance of the IMS, combined with wearable monitoring, in comparison to the intermittent paper-based EWS system. We calculated the time between consecutive observation sets performed within the 36 hours preceding a major adverse event. Similarly, for all patients who had no major adverse event, the time between each pair of observation sets was determined. The total monitoring time for patients in each trial phase using both bedside and telemetry monitoring systems was calculated. We determined both the time that patients were connected to a monitor and the time where data were actually acquired.

Post-hoc, to evaluate whether the change of the paper-based EWS system between both phases of the study had affected outcomes, we computed the proportion of observation sets which led to a clinical alert in each phase by the corresponding EWS system used at the time of the study.

### Data sources

Participants’ demographic information, time of discharge from the ward, and time of death were retrieved from the hospital’s patient administration system. Ward length-of-stay was calculated from the first observation set recorded on paper on the ward to the time of discharge. Time of admission to ICU from the ward was recorded from the Intensive Care clinical information system. Details of cardiac arrest calls were extracted from the clinical notes by research staff. All vital signs and associated early warning scores and times recorded electronically were automatically stored in the trial server. All observations and early warning scores recorded on paper were double-entered by separate research staff and reconciled into the trial database (14). Changes in ward practice were logged in the trial log. The hospital changed the paper-based EWS system (but not the escalation protocol) between the two phases.

### Statistical methods

We compared ward length-of-stay for each phase using Kaplan-Meier curves, (assigning individuals who died the “worst” outcome - i.e., right-censoring them at the longest recorded length-of-stay). We undertook time to event analyses for in-hospital mortality and ICU admissions between the two phases using the same methods. Confidence intervals for difference of two medians were computed also computed (15). Other binary outcome measures, such as cardiac arrest rates, were compared using Fisher’s exact tests. We compared the frequency of standard care observation sets during both phases of the study using Wilcoxon rank-sum tests. Confidence intervals for the difference of categorical (binary) measures were also estimated (16).

We note that the calculated *p* values were not corrected for multiple comparisons. For each endpoint we had complete data for analysis.

## RESULTS

Phase-I ran from May 2009 to June 2011. Phase-II ran from August 2011 to December 2013. 233 participants in phase-I and 250 participants in phase-II agreed to take part in the study, of which 200 and 207 participants took part, respectively (Figure 2). Participant demographic descriptors are detailed in Table 1. Participants were monitored with Visensia IMS (using either bedside or telemetry monitors) for 30.2% of their stay in phase-I and 58.2% in phase-II (Table 2, Supplemental Digital Content – Figure-A1). The most common reason for time without monitoring was participants choosing not to continue being monitored as they neared discharge. During monitoring, data gaps were most commonly due to equipment removal for patient care either temporarily (40.6% of logged gaps) or until the next research nurse check (23.3% of logged gaps). 12.7% of logged gaps were due to technical problems such as exhaustion of the batteries in the wearable monitors, malfunction of the wearable sensors and PDAs, power failures, or failures in the hospital wi-fi network. The median (IQR) length-of-stay of participants was similar in both phases: 8.9 (6.7–14.0) days in phase-I, and 9.0 (6.0–13.8) days in phase-II. Kaplan-Meier analysis (Supplemental Digital Content – Figure-A2) showed no statistical difference in the length-of-stay between both phases (log-rank test, *p*=0.327).

**Table 1.** Comparison of phase-I and phase-II participants’ demographics and clinical information.

|  | **Phase I**<br>**(n = 200)** | **Phase II**<br>**(n = 207)** |
| --- | --- | --- |
| Gender (male), n (%) | 111 (55.5) | 121 (58.5) |
| Age, median (IQR) | 63 (54 - 70) | 63 (51 - 69) |
| ASA grade, median (IQR) | 2 (2 - 2) | 2 (2 - 2) |
| Elective ICU, n (%) | 83 (41.5) | 75 (36.2) |
| Surgery type, n (%): |  |  |
| Whipples/bypass | 85 (42.5) | 73 (35.3) |
| Oesophagectomy | 39 (19.5) | 40 (19.3) |
| Hepatectomy | 34 (17.0) | 49 (23.7) |
| Gastrectomy | 23 (11.5) | 23 (11.1) |
| Splenectomy | 12 (6.0) | 9 (4.3) |
| Gastric-bypass | 7 (3.5) | 13 (6.3) |

**Table 2.** Monitoring time for study participants. We define one hour with data if at least 10 minutes of (continuously) data are available during every consecutive 60-minute window.

|  | Phase I (n = 200) | Phase II (n = 207) |
| --- | --- | --- |
| **Bedside monitor:** |  |  |
| Number of patients, n (%) | 105 (52.5) | 175 (84.5) |
| Cumulative hours monitored | 5018 | 11162 |
| Cumulative hours monitored with data | 2590 | 6012 |
| Proportion of hours monitored with data | 78.8 (40.3 - 99.3) | 76.9 (47.4 - 95.2) |
| **Telemetry monitor:** |  |  |
| Number of patients | 165 (82.5) | 198 (95.7) |
| cumulative hours monitored | 8825 | 15294 |
| Cumulative hours monitored with data | 7155 | 13058 |
| Proportion of hours monitored with data | 100.0 (77.6 - 100.0) | 100.0 (83.9 - 100.0) |
| **Proportion of time monitored (tablet + telemetry) during length of stay on the ward** | 30.2 (13.8 - 49.2) | 58.2 (33.1 - 75.2) |

**Figure 2.**
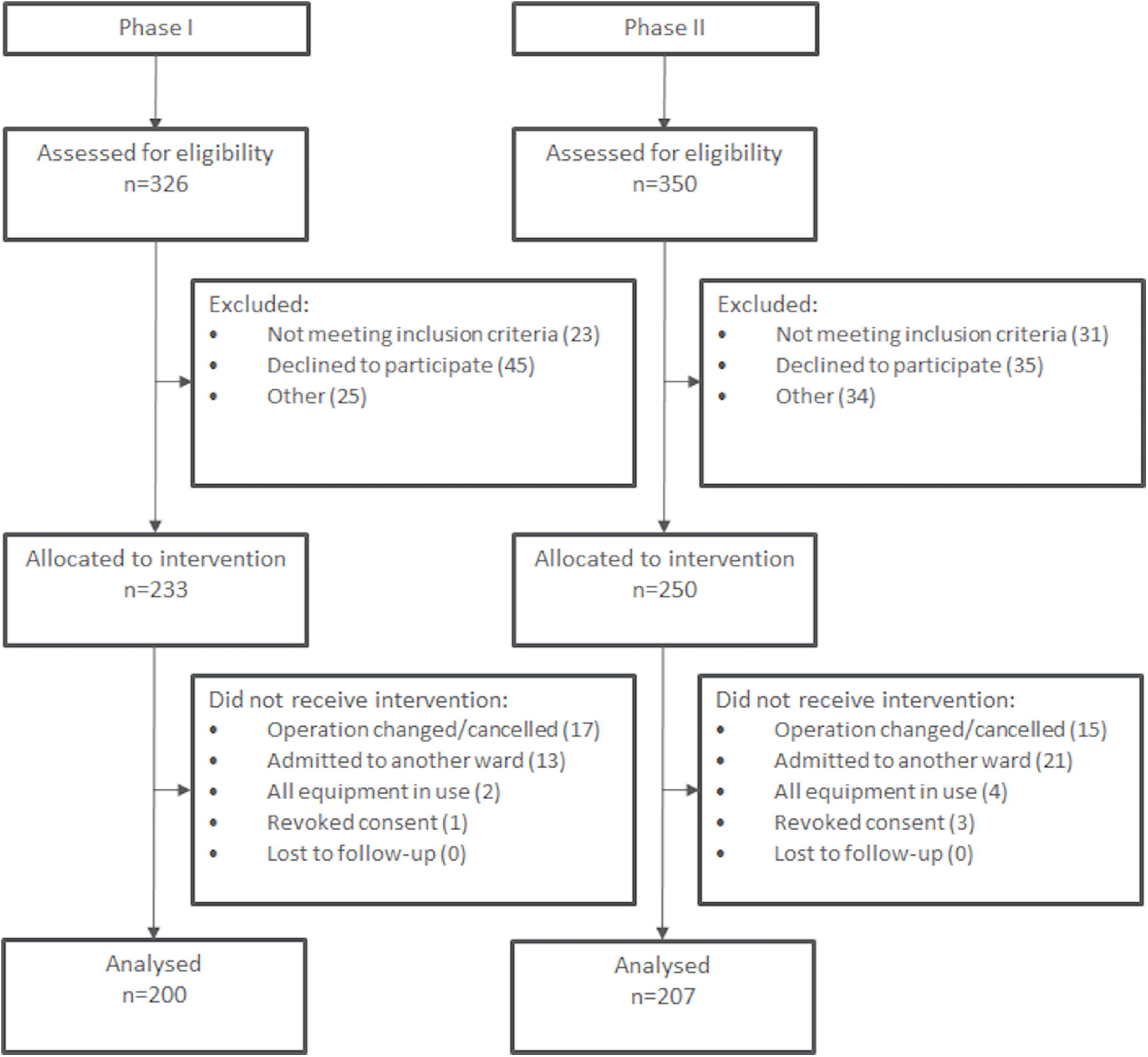
Consort diagram showing the flow of participants through each phase of the study.

Participants with adverse events had vital sign observations undertaken more frequently in the 36 hours preceding the event (the “critical period”) in phase-II in comparison to phase-I, with median (IQR) time to next observation in phase-I vs. phase-II of 1.0 (0.5-2.1) vs. 1.5 (0.8-2.5) hours (p<0.001). Participants with no adverse events had vital sign observations undertaken with similar frequency in both phases (Table 3).

**Table 3.**
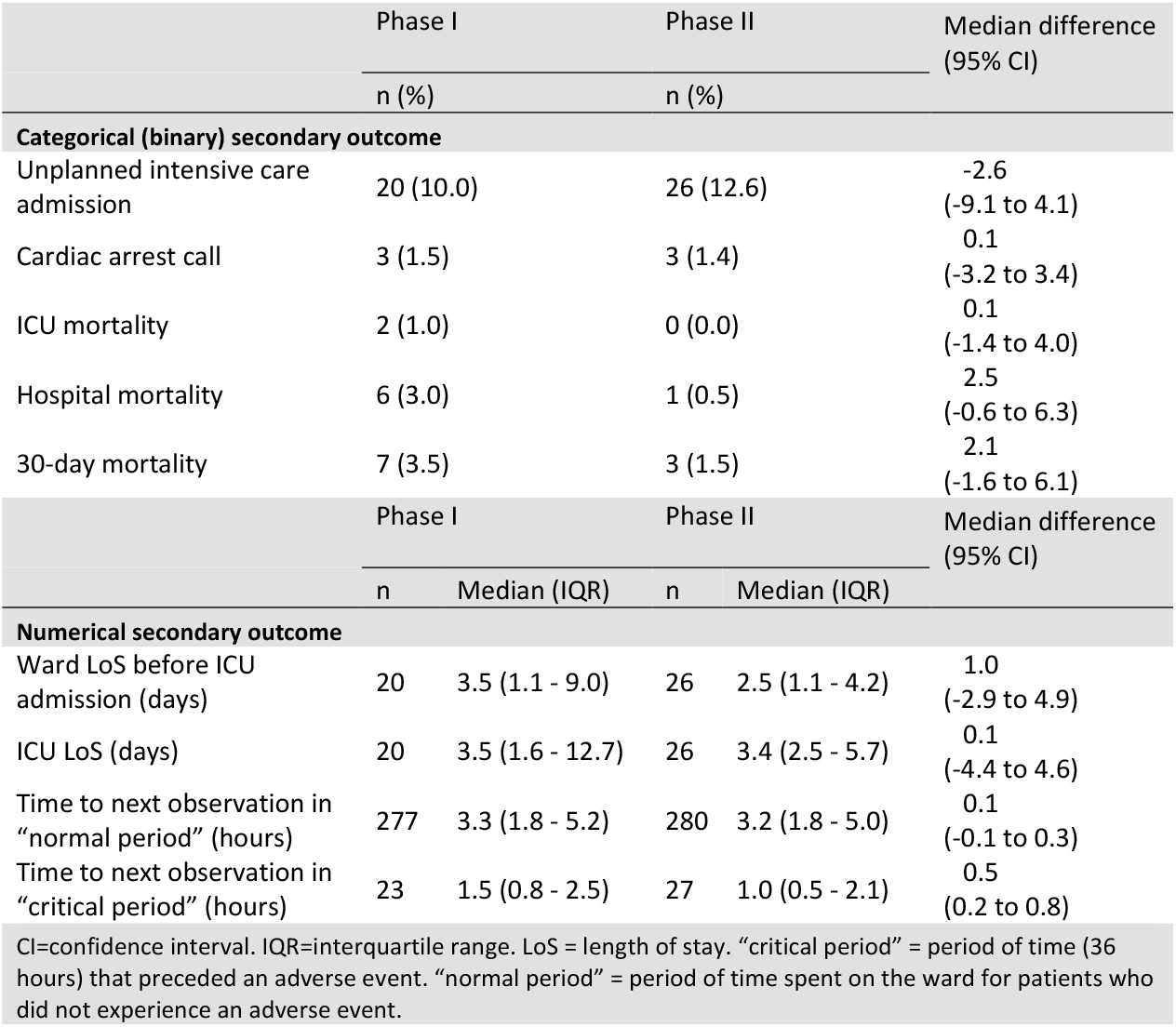
Comparison of the different secondary outcome measures.

In phase-I, six participants died before leaving the hospital. One participant died in phase-II (*p*=0.064) – Table 3. In phase-I, 20 participants had an unplanned admission to ICU (of whom three died whilst in hospital). In phase-II, 26 participants were admitted to ICU, all of which were subsequently discharged alive from the hospital. Unplanned admissions to ICU occurred at a median (IQR), 3.5 (1.2–9.0) days after admission to the ward post-operatively in phase-I, and 2.5 (1.2–4.3) days after admission to the ward in phase-II (*p*=0.327). The time spent by participants in ICU (for the unplanned ICU admissions) was similar in both phases (Supplemental Digital Content - Figure-A3).

There was no significant difference between the proportion observation sets which led to a clinical alert generated in both phases of the study for participants who either died or required ICU admission due to the paper-based EWS systems (the median, IQR, proportion of escalations generated by the paper-based early warning system used in phase-I was 5.6, 2.1-12.0%, and by that used in phase-II was 5.3, 2.2-11.4%, *p=*0.393).

## DISCUSSION

The addition of ambulatory continuous pulse oximetry and Visensia IMS to the standard paper-based early warning system did not reduce hospital length-of-stay following major upper-gastrointestinal surgery. However, implementing the system resulted in a change in care. In the 36 hours prior to an adverse event, patients had observations taken much more frequently. Six people died in hospital in phase-I. One participant died in phase-II. The one patient who died in phase-II was erroneously not monitored for 8.4 hours prior to a fatal cardiac arrest secondary to pulmonary embolism. All participants unexpectedly admitted to ICU in phase-II were discharged home alive, whereas two participants died before discharge following unanticipated admission in phase-I.

Our study is limited by its before-and-after design. We took this approach for several reasons. Ambulatory monitoring had never previously been attempted with a real-time computation of a risk score as part of the overall monitoring system. Phase-I gave us the opportunity to develop standard operating techniques to improve data capture and to check that the real-time risk score was appropriate for use with ambulatory monitoring in this patient population. The intervention was necessarily unblinded and staff would therefore tend to learn the combinations of vital signs that caused the real-time risk score to rise above 3.0 (alerting criterion) – potentially affecting their management of a randomised control group. We were concerned that in a constrained resource system, alerting using the real-time risk score in a randomised selection of participants at the same risk could disadvantage the care of the control group, present at the same time. Our study will allow the development of a future multi-site cluster-randomised study to answer many of the problems associated with this study design.

Before-and-after studies are primarily weakened by changes in clinical management or quality improvement interventions that typically occur on a hospital or ward level. In our study, for example, the organisation changed their paper-based early warning system between the two phases of the study. We carefully monitored for these confounders and observed that vital-sign monitoring and paper-based charting behaviour for those who did not have an event did not change between the two phases. The paper-based system in phase-II was not more sensitive to the participants with events than that in phase-I.

We wished to establish the effect of changing the monitoring and the visibility of that monitoring on patient care without changing the staffing required. We therefore did not provide an additional response team. However, ICU staff were always available should the ward staff require them.

Participants were monitored for 58.2% of their stay in phase-II of the study. This degree of monitoring improves considerably on using static monitoring alone (12), suggesting that ambulatory monitoring is a useful addition to post-operative patient care. However, it remains possible that with more unobtrusive monitoring, further gains can be made. We postulated that by improving early recognition of deterioration we would reduce length-of-stay. This did not occur. We had no data on which to base a likely reduction as our intervention was innovative. We chose what we thought would be a difference significant to participants (2-day reduction in length-of-stay). Improving recognition of deterioration only has the potential to improve outcomes in those who deteriorate in a manner where early deterioration will improve outcome. Around one in eight participants had a substantial deterioration (adverse event). Although participants will have deteriorated and received effective treatment without needing intensive care admission, the large majority of participants recovered uneventfully. As a result, the large majority of participants could not have their outcome positively affected by our intervention.

Our study is the first study to link real-time risk score estimation and alerting with ambulatory monitoring. Previous work has explored the deployment of real-time patient monitoring systems based on wireless technologies without evaluating prospectively the impact of these technologies on patient outcomes (17,18,19). In other work, the same IMS system has been associated with a reduction in the number and duration of periods of physiological instability in continuously-monitored non-ambulatory participants (i.e., confined to bed) in a post-surgical high-dependency environment (13). Taenzer et al. (20) had also conducted a before-and-after study where a pulse oximetry surveillance system (using single-parameter scoring criteria) resulted in a reduced need for ICU transfers. In our study, although the intervention did not alter ward length-of-stay, the results were equally encouraging. Patients who underwent serious deterioration had more nursing observation sets documented when monitored using the study system. This was not a feature of the system. When a participant’s physiology resulted in an alert, the icon on the large review screen flashed red until clinical staff acknowledged the alert. However, extra observation sets were not suggested. The system therefore resulted in clinical staff recording more vital-sign sets on deteriorating patients. The frequency of vital-sign recording was unchanged in participants who did not undergo a substantial deterioration. Hence, implementing the system resulted in improved focus of care on those at risk of serious deterioration. Increasing the documented monitoring of patients who are at risk of substantial deterioration is a change in behaviour that must underlie early recognition of these patients. It is the first step to improving outcomes. The study was not powered to detect a mortality benefit, but the difference in mortality seen between the two phases is encouraging.

## CONCLUSIONS

Combining real-time alerting using a multi-dimensional model of normality with ambulatory monitoring in high-risk post-surgical patients improves recognition and management of deteriorating patients without increasing the workload of clinical staff in those patients who do not deteriorate.

## Data Availability

N/A

## Ethical approval and Trial Registration

This study was approved by the Oxford Research Ethics Committee OxREC No: 08/H0607/79.

## Acknowledgements

We wish to thank Breda Lynch, Theresa Saunders and Deborah Evans for their work on the study, Jacqueline Birks for undertaking statistical analysis and Julie Darbyshire for proof-reading the study. The study could not have been undertaken without the support of the clinical staff on the upper-gastrointestinal ward. Most importantly, we would like to thank the participants of the study for their commitment to research and particularly to wearing additional ambulatory monitoring post-operatively.

## Supplemental Digital Content

**Table-A1**. Study participants’ exclusion criteria.

**Table-A2**. Paper-based early warning scoring systems in use during the study.

**Figure A1.**
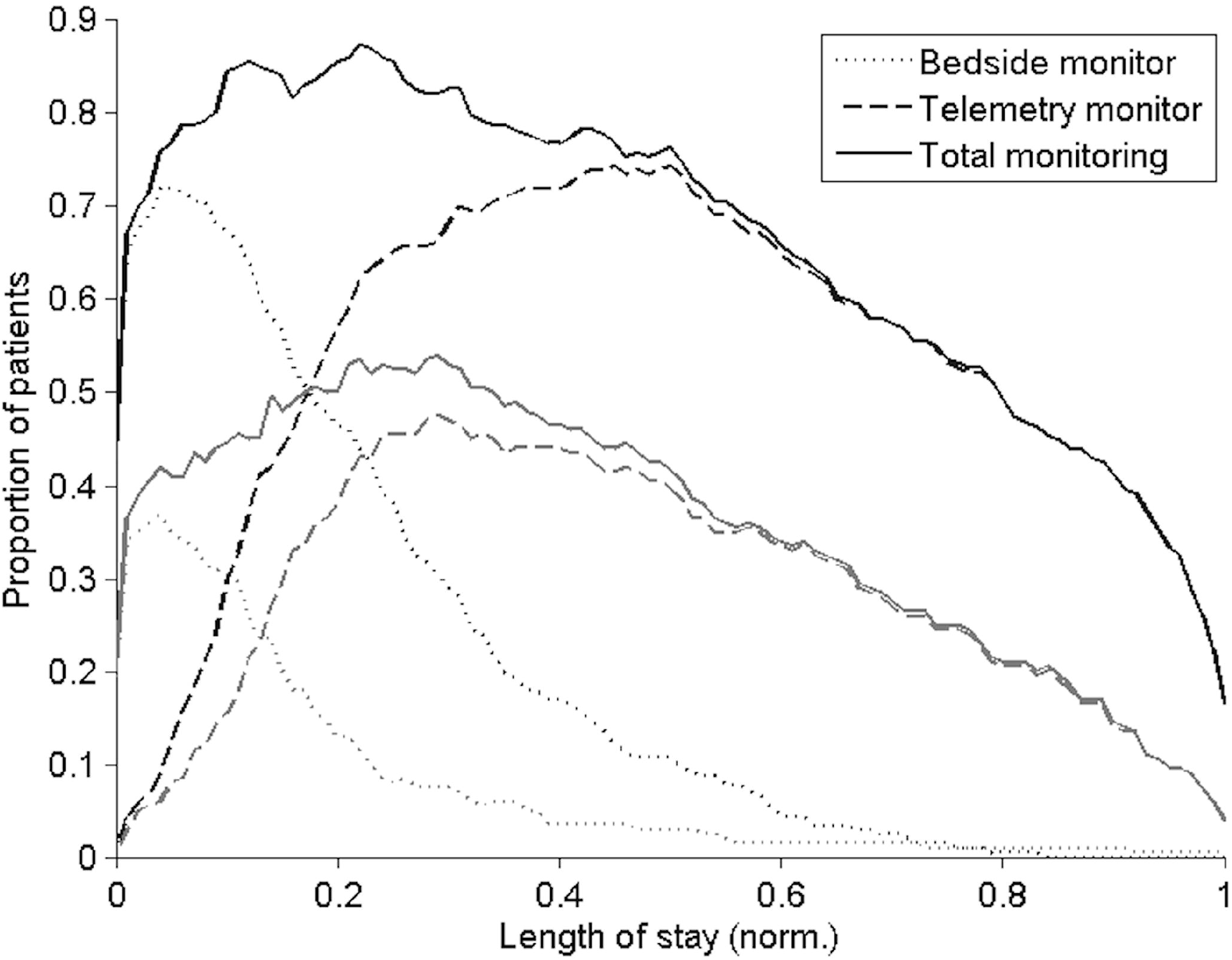
Proportion of participants monitored (using both bedside and telemetry monitors) over their length-of-stay on the ward (normalised to a scale 0 to 1): darker lines correspond to phase-II, and lighter lines correspond to phase-I.

**Figure A2.**
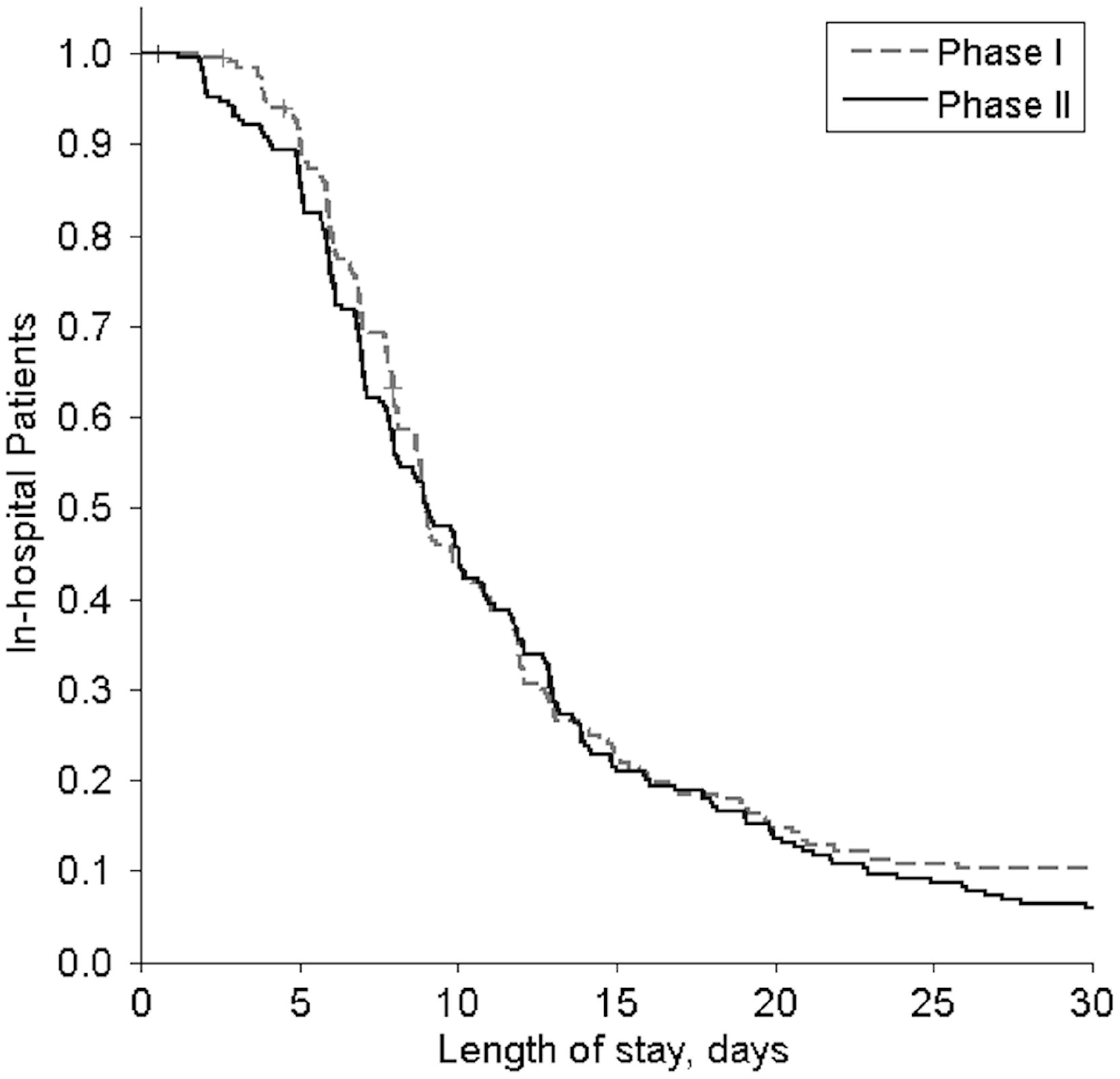
Kaplan-Meier curves of ward length-of-stay (LoS) after major upper gastrointestinal surgery. Phase-II was not associated with a significantly shorter LoS (log-rank test, *p* = 0.327).

**Figure A3.**
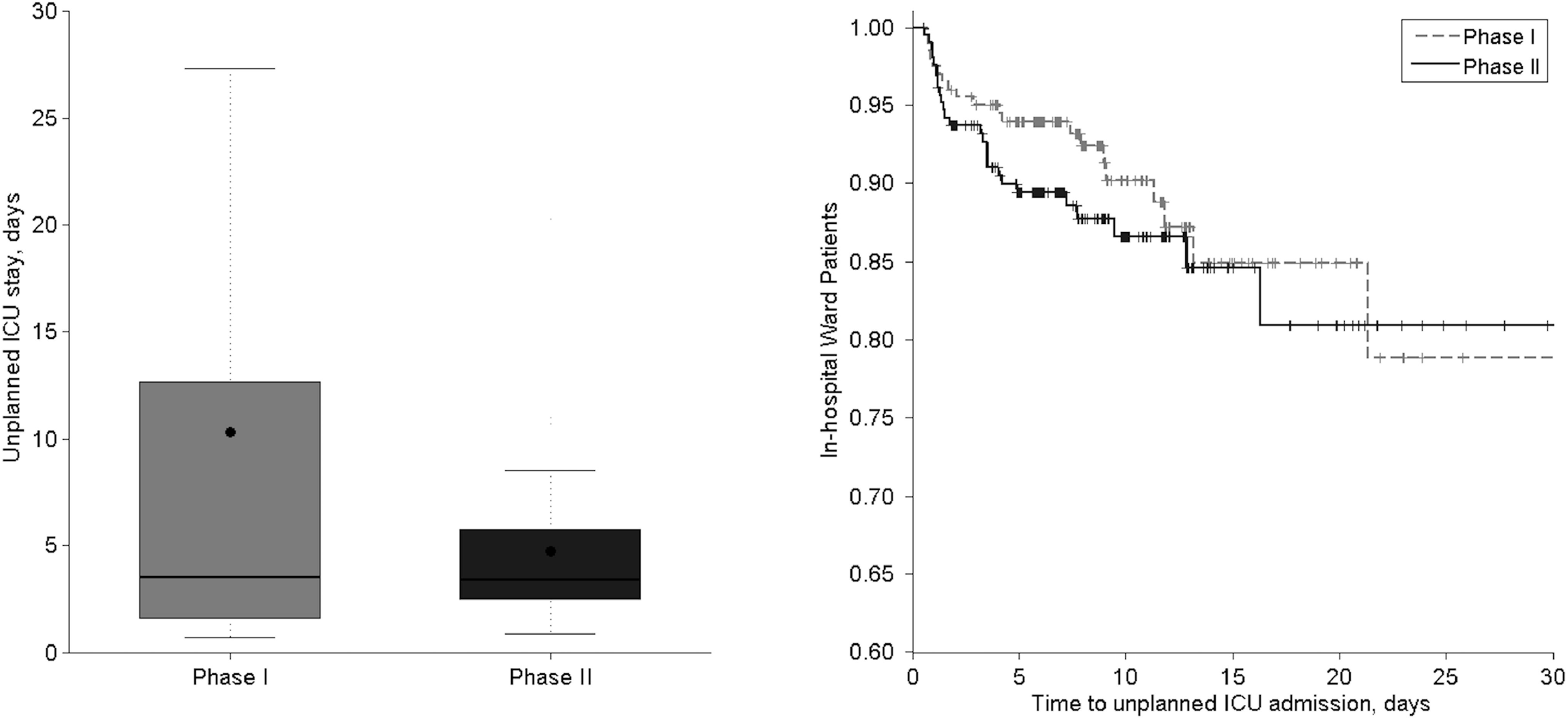
(Left) Box-Plot of length-of-stay in intensive care, after an unplanned admission to intensive care, for both phases (*p* = 0.797); black dots correspond to the mean values. (Right) Kaplan-Meier curves of ward length-of-stay before an unplanned admission to intensive care, also called time to unplanned ICU admission (log-rank test, *p* = 0.431).

