## Supplementary material for "Early detection of physiological deterioration in post-surgical patients using wearable technology combined with an integrated monitoring system: a pre- and post-interventional study": Table A1

**Supplemental Digital Content**

**Table A1.** Study participants’ exclusion criteria.

Exclusion criteria of both phases of the study:

- Participants under 16 years of age
- Pregnant Women
- Participants unable to wear the required equipment
- Participant who could not understand written English, for whom no translator could be found.
