## Supplementary material for "Early detection of physiological deterioration in post-surgical patients using wearable technology combined with an integrated monitoring system: a pre- and post-interventional study": Table A2

**Supplemental Digital Content**

**Table A1.** Paper-based early warning scoring systems in use during the study.

| Phase I paper-based early warning score system  A score of 4 or more triggers a review of the participant. | | | | | | | |
| --- | --- | --- | --- | --- | --- | --- | --- |
|  | Score | | | | | | |
| Variable | 3 | 2 | 1 | 0 | 1 | 2 | 3 |
| Heart rate |  | ≤ 40 | 41 - 50 | 51 - 100 | 101 - 110 | 111 - 129 | ≥ 130 |
| Resp. rate | ≤ 8 |  |  | 9 - 18 | 19 - 24 | 25 - 29 | ≥ 30 |
| Temperature |  | ≤ 35.0 |  | 35.1-37.9 |  |  | ≥ 38.0 |
| Systolic BP | ≤ 90 | 91 - 99 |  | 100 - 179 |  |  | ≥ 180 |
| SpO_2_ | ≤ 92 |  |  | ≥ 93 |  |  |  |
| AVPU scale |  |  |  | A | V | P | U |
| Phase II paper-based early warning score system  A score of 3 or more triggers a review of the participant. | | | | | | | |
|  | Score | | | | | | |
| Variable | 3 | 2 | 1 | 0 | 1 | 2 | 3 |
| Heart rate | ≤ 42 | 43 - 49 | 50 - 53 | 54 - 104 | 105 - 112 | 113 - 127 | ≥ 128 |
| Resp. rate | ≤ 8 | 9 – 11 | 12 - 13 | 14 – 25 | 26 - 28 | 26 - 33 | ≥ 34 |
| Temperature | ≤ 35.4 |  | 35.5-35.9 | 36.0-37.3 | 37.4-38.3 |  | ≥ 38.4 |
| Systolic BP | ≤ 85 | 86 – 96 | 97 - 101 | 102 - 154 | 155 - 164 | 165 - 184 | ≥ 185 |
| SpO_2_ | ≤ 84 | 85 – 90 | 91 - 93 | ≥ 94 |  |  |  |
| AVPU scale |  |  |  | A | V |  | P, U |
